# Impact estimation on COVID-19 infections following school reopening in September 2020 in Italy

**DOI:** 10.1101/2021.06.11.21258733

**Authors:** Livio Fenga, Massimo Galli

**Author notes:** Corresponding author: Livio Fenga.

## Abstract

**Background:** Since its outbreak, CoViD-19 (formally known as 2019-nCoV) has been triggering many questions among public authorities, social organisms and school officials, as to when students should be allowed to return to school. Such a decision is critical and must take into account, other than its beneficial effects, also those associated with an increased exposition of the students to the virus, which, as a result, might spread at a faster rate. To date, in Italy, a few studies have rigorously investigated the correlation between school reopening and number of people tested positive to CoViD-19. Therefore, this paper aims to provide an assessment of such an impact as well as to illustrate the methodology followed.

**Methods:** Official daily data on the cumulative number of people tested positive to CoViD-19 – in conjunction with external information accounting for the different points in time schools reopened in the various Italian regions – have been employed to build a stochastic model of the type Seasonal Autoregressive Moving Average embodying external information.

**Results:** There was a statistically significant increase in the number of positive cases in all the Italian regions related to schools reopening. Such an increase occurred, in average, about 18.9 days after the schools have been reopened. Schools reopening have been significantly contributed to the diffusion of the pandemic, with an overall estimated impact of about 228,724 positive cases.

**Conclusions:** The results suggest the need for strict control of all in-school activities. This could be done by using, to a variable extent, all the non-pharmaceutical interventions available, such as limited access to school spaces, no overlapping practices between different sports in the same space, universal masking, bubble-size classroom. However, in many cases, such measures might not be a viable option, at least in the short run, nor be reasonably applicable. Therefore, whenever the established safety criteria could not be met, school buildings should remain closed.

**Key Messages:**

- Due to CoViD-19 pandemic, physical school attendance is at the center of an intense political and social debates;
- schools reopening decision should be based on reliable and well maintained data-sets;
- in the lack of quality data, it is advisable to use a portion of them, to reduce uncertainty.

## Introduction

Especially during epidemics, morbidity and mortality due to secondary bacterial infections can be substantially increased when specific occasions of congregation – e.g. physical school attendance, eating in bars and restaurants, attending sporting events – are permitted by public authorities. In such a scenario, potentially dangerous relaxations of some of the countermeasures – purposely taken to contain the spread of the virus – are unfortunately un-avoidable. This is the case of CoViD-19 pandemic, which, among others, has presented unprecedented challenges to the Italian education system, which led to the decision of closing most of the schools between March and June 2020. Such a decision triggered many protests with some of them, as reported by all the major national media, resulting in clashes and other social disorders as a consequence of violent and anti–social behaviors. The crisis is still ongoing and this is a delicate matter, given its potential ability to jeopardize the social and democratic fabric of a country^1 2^. With the school closed, potential educational risks have been highlighted by many experts (e.g. psychologists, professional educators, school officials). In more details, it is noted that hardly ever do students of any age group can fulfill their educational needs in remote learning modes the same way they do under in–person set-ups. In addition, remote learning can have adverse effects on young people in terms of their future social abilities^3^, promote inequalities and reduce the effectiveness of interventions against the risks of suicidality^4^. From a broader perspective, keeping the school open has generally a positive impact on parents’ working life, beyond educating children. In fact, families are relieved from all the duties generally fulfilled by schools, e.g. a safe environment, provision of meals, in-school health services. On September 2020, after a vigorous public debate, a controversial decision to reopen the schools was finally taken by the Italian government. This paper focuses on the impact generated by such a determination on the diffusion of CoViD– 19. The analysis has been carried out at regional level, using official data, as detailed in Section 4. Given the scarcity of school-specific information, population-wide time series – related to the cumulative number of people tested positives (henceforth positive cases or simply positives), stratified at a regional level – have been used. The statistical procedure adopted is based on a stochastic model of the type S-ARIMA– REG (short for Seasonal Auto Regressive Integrated Moving Average with external REGressor), which is designed to model exogenous information, that is, in our case, the different dates on which schools reopened (according to the rules established by the regional public authorities). As it will showed, this model proved to be effective in adequately capturing the effect of school reopening on the dynamical behavior of the time series of the cumulative number of positives.

## Literature review

The effects of learn-at-home strategies in flattening the CoViD-19 pandemic curve, have been studied worldwide from various points of view and using different approaches. A comprehensive, systematic review and meta-analysis of 2178 articles, carried out in^5^, draws (at least) two important conclusions: the lower risks of contracting the infection associated to young people and the overall poor quality of a big chunk of the studies considered. The latter is a particularly striking message as it might be intended as a caveat on some of the results today available in literature. In this regard, a better insight on many of them can be gained by means of the results of a mass seroepidemiological screening, performed in a zone of Italy characterized by unrestricted viral circulation (Castiglione d’Adda), presented in^6^. With that said, there are published and non peer-reviewed articles claiming the limited or even the absence of a significant direct correlation between physical attendance in schools and spread of the virus. In general, such a conclusion is grounded on the fact that children have lower susceptibility to CoViD-19 compared with adults^7^. In this regard, a conservative approach should be followed by taking into consideration that an apparent lower incidence in children might be induced by facts such as reduced exposure and methodological issues, including lower testing^8^. In addition to that, it has to be said that hardly ever do children show obvious symptoms and that school settings often envision small classes and extensive hygiene measures. In a research study focusing on school activities in Italy^9^, it is outlined how the spread of the virus among students occur outside the school buildings, e.g. as a consequence of insufficient transportation or unauthorized social gatherings. However, it seems to be unclear whether such a conclusion is adequately supported by reliable data sources. Intra-class transmission has been found to be a rare event also in a preliminary study^10^ whereas in^11^ no evidence of association between schools and CoViD-19 second wave have been found in Italy. While the former study seems to lack a thorough explanation of the method applied, in the latter the authors explain the two-fold approach followed: a cross sectional and prospective cohort study. However, part of their research questions have been supported on a data sample of limited size, e.g. the Veneto region or one of its province (Verona). In addition, it is not entirely clear how the cross–correlation functions have been estimated (e.g. it might be advisable to use prewhitening techniques^12^ in order to draw more informative conclusions). A prospective, cross-sectional study^13^ – carried out in England based on official data – concluded that only “very few cases” of CoViD-19 are attributable to schools opening. In more details, a child has been detected as the potential source of infection in only 29% of child cases and 17% of staff cases. Other results for England, reported in^14^, indicate not significant transmission rates in primary schools, before the new CoViD-19 variants started spreading. The impact of school closure in Ontario, Canada, has been assessed in^15^: the results obtained indicate that school closure had a limited impact on the attack rates. If, on one hand, these studies can be reassuring and have positive effects on the general mood, on the other hand – being based on the assumption of a comparatively small proportion of CoViD-19 cases in children – there is a substantial risk of making wrong inferences on the CoViD-19’s transmission capabilities among children in school places. Consistently, many articles warn about the dangers associated with schools reopening: in an extensive study^16^ a significant correlation between school closures and the reduction of the reproduction number *R*_*t*_ has been found for different time windows and across many countries. A study^17^ carried out in Israel found that, compared with the closing period, the proportion of infected children increased from 19.8% to 40.9%. In the same direction goes the excellent work^18^, where a clear patio–temporal correlation between school and (delayed) increase in the overall contagion has been showed shown for Italy. The Italian case has been also thoroughly examined within a Bayesian framework in^19^. In their work, the authors found in 15 out of 21 Italian regions – after an average delay of 16.6 days – a change in the rate of growth of the cumulative number of positive cases. Finally, the association between statewide school closure and COVID-19 incidence and mortality has been considered for the US in^20^. The authors found a significant relation between the reduction in both number of positive cases and mortality rate and school closures, even though other concurrent non pharmaceutical actions might have played a role in such a reduction.

## Method

The mathematical methods generally used for description, prediction and simulation purposes in the case of CoViD-19 are those typically used for other epidemic events (e.g. Ebola, Zika, MERS-CoV). For a review of the most used approaches, the reader is referred to21 22 23. As above mentioned, the results provided in this article are obtained within the framework of the time series analysis. In particular, a stochastic model of the type S-ARIMA–REG has been employed (to the best of the authors’ knowledge, this is the first time such a method has been applied to address research questions similar to that investigated in this paper). Time dependent data have been extensively employed in epidemiology^24 25 26 27 28 29 37^ and modeled by a variety of stochastic models, including S-ARIMA. Introduced in 1970 by Box and Jenkins^33^, these type of models have been successfully used in many fields of research, including epidemiology, see e.g.^24 29 16^. A S-ARIMA-REG model takes the form of a time t, *t* ∈ ℤ^+^, indexed difference equation which can be expressed as

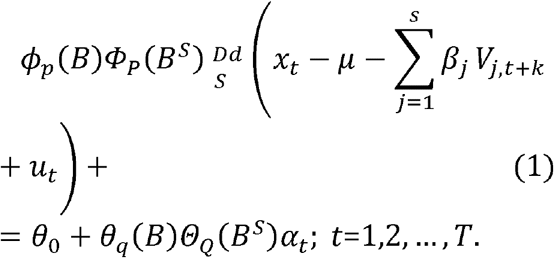

Denoting with *B, d*, and *D* the backward shift operator and the non-seasonal and seasonal difference operator, respectively, defining ^*d*^ =1 − *B*^*d*^ and ^*D*^ =1 − *B*^*D*^, we have *ϕ*_*p*_(*B*) =1 − *ϕ*_1_*B* − *ϕ*_2_*B*^2^−.… −*ϕ*_*p*_*B*^*p*^, *θ*_*q*_ (*B*) =1 −*θ*_1_B −*θ*_2_*B*^2^−. … − *θ*_*q*_*B*^*p*^, *Φ*_*P*_(*B*^*S*^) = 1− *Φ*_1_*B*^*S*^ − *Φ*_2_*B*^2*S*^−.. .. −*Φ*_*P*_*B*^*PS*^, and *Θ*_*Q*_(*B*^*S*^) =1 − *Θ*_1_*B*^*S*^ − *Θ*_2_*B*^2*S*^−. … −*Θ*_*q*_ *B*^*QS*^. Here, *ϕ,θ, Φ, Θ*, respectively, denote the non-seasonal autoregressive and moving average parameters and the seasonal autoregressive and moving average parameters. Finally, *α*_*t*_ is a 0—mean white noise with finite variance *σ*^2^ whereas the estimation algorithm adopted in this paper is of the type Maximum Likelihood. Exogenous information is captured by the matrix *V*_*j,t*+*k*_, being *k* ∈ ℤ^+^, weighted by the parameter vector *β*_*j*_. In our setup, however, the schools reopening date is the only external input – and thus the matrix *V*_*j,t*+*k*_ collapses to a simple 0 − 1 dummy vector, denoted by the symbol *V*_*t*+*k*_, weighted by the scalar parameter *β*_*j*_. In more details, the vector *V*_*t*+*k*_ takes the value 0 before the school opening plus a lag *k* and 1 afterwords. In such a way, the *β* parameter expresses a steady mean variation – which is known also with the term “level shift” – from the time the dummy variable transitions from the state 0 to 1 onward. The integer *k* plays a key role in our analysis, as it captures the delay at which the greatest effect on cumulative positives cases is detected. Due to its efficient design, Equation 1 can describe two scenarios: one assuming that the intervention (schools opening) has never happened – and one which “acknowledges” it. The former, also known as a counterfactual or hypothetical scenario, is built by simply subtracting the estimated impact *β* to the series of cumulative positives located from *t*_0_ +*k* onward, with *t*_0_ being the school starting date. In symbols *x*_*t*_ − *β*; *t* = *t*_0_, *t*_0+1_,…,*T* − 1, *T*, being *T* the last observation in the observed sample and *x*_*t*_ the variable of interest. Due to the observed systematic reduction of the number of tests administrated during the weekends, a S-ARIMA structure – designed to account for periodic components embedded in the underlying stochastic process – has been chosen over the simpler ARIMA model (where no periodicity is accounted for).

The algorithm employed to generate the results, presented in the next Section 4, iteratively maximizes the explanation capabilities of the vector *V*_*t*+*k*_ (Equation 1) through a grid search approach, performed over a set of reasonable tentative delays *k*_1_, *k*_2_,…, *k*_*K*_. In practice, model (1) is re-estimated *K* − *k* + 1 times (the models have been always checked for both stationary and invertibility conditions^2^) and the final S-ARIMA-REG structure selected using the Minimum Akaike Information Criterion Estimate procedure^33 34^, being the grid search exhaustively examined in the interval [7; 60]. Recalling *t*_0_ to be the school starting date, for each region the algorithm extracts the delay *t*_0_+ *k*^*^ at which the maximum impact - in terms of positive cases – is generated by the schools opening. Such an assessment, is based on the largest statistical significance, expressed by the value of the *t*-test, associated to the (*K* − *k* + 1) *β* parameters recursively estimated. In symbols: 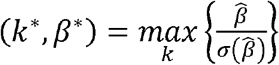 under 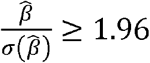 (i.e. the 95% confidence interval for which *β* ≠ 0). In practice, for each region the algorithm returns the sequence of 60 − 7 + 1= 54 outcomes of the *t*-test, whose maximum is taken to select the value of *k*, i.e. *k*^*^, as the day number, following the relaxation of the schools closure (*t*_0_), where the impact of such a measure is maximized. For example, Figure 1 portrays the sequence of t-values (one for each day between day 7 and day 60 after the schools starting dates) related to the Trento region. The peak located on October 9, 2020 indicates that the greatest impact has occurred *k*^*^ = 37 days (*t*^*^ = 11.1) after the schools have been reopened. Once the time delay has been established, the estimated *β* parameter associated to the dummy variable *V*_*t*+37_ accounts for the number of positive cases attributable to the event schools opening (*t*_0_) shifted by *k*^*^ = 37 days, which in this case is equal to 256. In practice, the vector *V*_*t*+*k*\*_; *k*\* = 37 is simply a sequence of 0s between February 24 and October 8 and of 1s from October 9 onward. Being sampled–based, the estimation of the number of positives *β*^*^ generated by (1) has been extrapolated, for each region, to population scale through a suitable inference procedure^35 27^. In practice, the following multiplying factor, i.e.

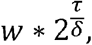

with

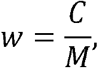

has been built, so that the final number of positives at a population level is given by 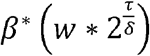. Here, *w* is the ratio between the current positive cases (*C*) and the number of deaths (*M*), *τ* the Covid–19 average doubling time (i.e. the average span of time needed for the virus to double the cases) and *δ* the average time needed for an infected person to die. While *δ* has been kept fixed and set to 6.2 (see^36^), the parameter *τ* has been empirically estimated for each region, using as a benchmark the number of positives recorded in the first school day and counting the number of days needed for this number to double. The number of deaths *M* are also available at the same official web address above given.

**Fig. 1.**
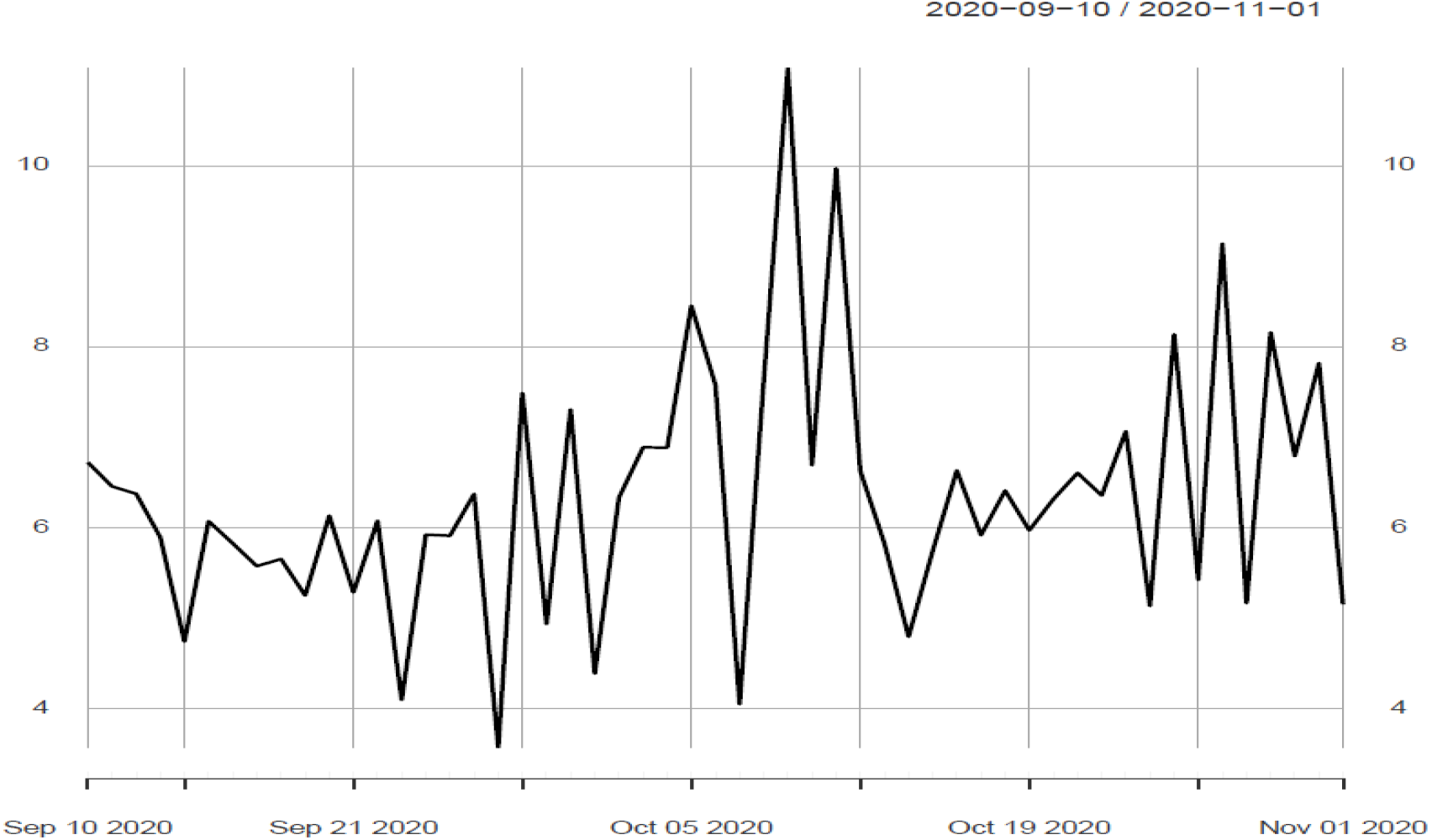
Outcome sequence of the *t*–*test* associated to the parameter *β* (i.e. the estimated additional number of positive cases) for the Trento region. The highest value *t*^*^ is reached on October 10, 2020

## Results

The data used in this paper are made available by the Italian Civil Protection Department and publicly accessible, free of charge, at the following web-address: https://github.com/pcm-dpc (file name: “dpc-covid19-ita-regioni-20210211.csv”). The variable of interest, stored in column 11 and labeled “Totale Positivi” (i.e. cumulative number of positives), consists of 407 daily data points collected at a regional level during the period February 24^*th*^ 2020 - April, 5^*th*^ 2021. Even though the official number of Italian regions is 20, one special administrative region (called Trentino Alto Adige) is considered as two separate subregions: Trento and Bolzano. Table 1 shows the results broken down for geographical distribution and each Italian region (respectively listed in the first and second column) in terms of the estimated number of positives cases attributable to schools reopening, reported in column 5. The statistical significance of these estimations is reported in column 6, whereas column 3 stores the schools starting dates. Finally, column 4 accounts for the number of days passed between the first school day and the day where the *t*-value reaches its maximum value. In almost all the cases – saved for Liguria and Veneto where a six days periodicity has been determined – the underlying seasonality chosen is always of 7 days. Such a periodic fluctuation in the data should not be associated with any epidemiological reasons being related to the screening procedures, which typically slow down on weekends. The estimated overall impact of schools reopening is quantified in around 227,724 positives whereas the mean time delay (henceforth denoted with the symbol *µ*_*D*_) is of about 19 days, with standard deviation (henceforth denoted with the symbol *σ*_*D*_) of around 7.5 days. Such estimates are consistent with what found in literature, see, e.g.. Schools reopening has been estimated to generate the greatest in the following regions: Lombardy, Piedmont (northern regions), Lazio and Tuscany (center regions), Campania and Apulia (southern regions). Those cases account for almost 50% of the estimated total impact. Lombardy is the most impacted region, with an estimated number of schools-related positives of more than 44,780 cases (delay time *k*^*^ = 30 days), whereas Calabria, Trento, Molise and Valle d’Aosta, are under the 300 positives threshold. Basilicata is the only region where the estimated model exhibits a not completely satisfactorily *t*-value (*t*^*^ = 1.8). The reason of that is probably linked to the high degree of roughness showed by the time series in the time window of interest, which might have resulted in a problematic convergence of the estimation algorithm. As for the time delay, for many regions our results are comparable to those reported in. In average, the values of *k*^*^ tend to decrease from north (*µ*_*D*_ = 23 and *σ*_*D*_ = 7.9) to center (*µ*_*D*_ = 20.2 and *σ*_*D*_ = .5) and south (*µ*_*D*_ = 14 and *σ*_*D*_ = 6.6).

**Table 1.** Impact of schools reopening in terms of positive cases (fifth column) broken down for geographic distribution and Italian regions (first and second columns). School starting dates are reported in the third column whereas the fourth one accounts for the number of days passed between the schools starting day and the effect in terms of positive cases (encoded in the vector *V*_*t*+*k*_) attributable to schools reopening. The last column 6 stores the greatest *t* values (*t*^⍰^). See text for details

| Geogr.<br>distrib. | Region<br>name | Schools<br>starting dates | Days to<br>max impact | School-related<br>positives | Statistical<br>significance |
| --- | --- | --- | --- | --- | --- |
| North | Piedmont | 09/14 | 27 | 16448 | 3.9 |
|  | Valle d'Aosta | 09/14 | 11 | 57 | 5.9 |
|  | Lombardy | 09/14 | 30 | 44780 | 2.8 |
|  | Bolzano | 09/07 | 27 | 800 | 6.6 |
|  | Trento | 09/03 | 37 | 256 | 11.1 |
|  | Friuli-Venezia-Giulia | 09/14 | 19 | 3644 | 6.3 |
|  | Liguria | 09/14 | 20 | 8504 | 7.8 |
|  | Emilia Romagna | 09/14 | 18 | 13697 | 2.6 |
|  | Veneto | 09/14 | 18 | 12917 | 6.8 |
| Center | Lazio | 09/14 | 21 | 23584 | 4.0 |
|  | Marche | 09/14 | 20 | 7011 | 6.0 |
|  | Toscana | 09/15 | 20 | 14846 | 4.1 |
|  | Umbria | 09/14 | 20 | 1292 | 3.3 |
|  | Abruzzo | 09/22 | 12 | 1228 | 22.3 |
| South | Basilicata | 09/24 | 9 | 1239 | 1.8 |
|  | Calabria | 09/24 | 9 | 271 | 2.9 |
|  | Campania | 09/24 | 8 | 37545 | 2.1 |
|  | Molise | 09/14 | 22 | 100 | 2.9 |
|  | Apulia | 09/24 | 9 | 24498 | 1.9 |
|  | Sicily | 09/21 | 19 | 11583 | 5.8 |
|  | Sardinia | 09/22 | 22 | 8784 | 4.0 |

## Discussion

The results obtained, clearly indicate the existence of a range of factors explaining the second wave of September in Italy. Our opinion is that those factors should include schools reopening, whose correlation with the rise of the epidemic curve has been statistically proven. These results call for a strict control of all the activities carried out in the school buildings. Such a goal can be achieved by using, to a variable extent, all the non pharmaceutical interventions available, such as limited access to school spaces, no overlapping practices between different sports in the same space, universal masking, bubble-size classroom, extensive hygiene. Reopening schools in a staged fashion – e.g. by year groups or location (e.g. rural or urban) – is thus an option, as proposed in^38^. However, it is a fact that in many cases, such measures might not be a viable option, at least in the short run, nor be reasonably applicable. Therefore, whenever the established safety criteria could not be met, these school buildings should remain closed. Lack of reliable information and intrinsic limitations of the S-ARIMA-REG scheme, prevented us from designing a more complex model, e.g. able to test and discriminate among different triggering factors. With that said, we believe that the methodological approach chosen has one main advantage over the methods based on multiple data sources: we used one single variable (the cumulative positives cases) and one auxiliary information (the schools starting date). As a result of that, the amount of uncertainty surrounding the whole analysis has been drastically reduced. In addition, the theoretical framework chosen makes possible the extraction of useful information – including counterfactual scenarios, as mentioned in Section 3 – on the dynamical behavior of the diseases and an easier interpretation of the results.

## Data Availability

n/a

https://github.com/pcm-dpc/COVID-19/tree/master/dati-regioni

## Notes

### Competing Interest Statement

The authors have declared no competing interest.

### Clinical Trial

not applic

